# Using extracted data from Kaplan-Meier curves to compare COVID-19 vaccine efficacy

**DOI:** 10.1101/2023.04.19.23288799

**Authors:** Amarit Tansawet, Pawin Numthavaj, Songporn Oranratnachai, Grace Filbertine, Sasisopin Kiertiburanakul, Gareth J McKay, John Attia, Ammarin Thakkinstian

**Author notes:** **Corresponding author Amarit Tansawet**, Department of Surgery, Faculty of Medicine Vajira Hospital, Navamindradhiraj University, 681 Samsen Road, Dusit, Bangkok, Thailand 10300.

## Abstract

Although various COVID-19 vaccines have shown efficacy against placebo in randomized clinical trials, no head-to-head comparisons are yet available. This study aims to compare the efficacy of available COVID-19 vaccines. Vaccine trials searched in May 2021 were included. Data were extracted from Kaplan-Meier (KM) curves using the WebPlotDigitizer program for the individual participant (IP) data simulation. A mixed-effect acceleration failure model with log-logistic and Weibull distributions was used to estimate relative effects for individual vaccines as well as grouped by class: inactivated virus, mRNA, and viral vector. Primary studies were considered as the random effect in the model. Hazard ratios (HR) were estimated and compared across vaccine groups. All vaccines were efficacious in lowering symptomatic infection compared to placebo. CoronaVac, Ad26.COV2.S, ChAdOx1 nCoV-19, rAd26/rAd5, WIV04, HB02, and BNT162b2 showed 7.61 (4.50, 12.87), 6.77 (4.08, 11.24), 5.01 (2.93, 8.57), 4.50 (2.52, 8.01), 3.90 (2.04, 7.45), 3.18 (1.62, 6.21), and 2.15 (1.22, 3.78) times significantly higher risk of infection than mRNA-1273. mRNA vaccines were the most efficacious vaccine group compared to inactivated virus and viral vectors with HRs (95% CI) of 0.27 (0.20, 0.37) and 0.28 (0.21, 0.37), respectively. Although all vaccines showed significant protection compared to no vaccination. mRNA vaccines, including mRNA-1273 and BNT162b2, showed the highest efficacy in preventing symptomatic COVID-19 infection. Simulated IP data from the KM curve might allow treatment comparison when there is no primary study comparing active treatments.

## 1. Introduction

The severe acute respiratory syndrome coronavirus 2 (SARS-CoV-2) has infected over 170 million people, caused over 3.7 million deaths (as of 1^st^ June 2021) (World Health Organization, 2021), and ignited global vaccine research. Various COVID-19 vaccines have been developed and tested in field trials, and are being used in vaccination programs worldwide. Current vaccines can be grouped according to their class: inactivated coronavirus (e.g., CoronaVac, BBIBP-CorV, and Covaxin), viral vector (e.g., ChAdOx1 nCoV-19, Ad26.COV2.S, and rAd26/rAd5), mRNA (e.g., mRNA-1273 and BNT162b2), and subunit vaccine (e.g., NVX-CoV2373). The efficacy of inactivated or viral vector vaccines has been reported to range between 50.7% and 91.5% (Logunov et al., 2021; Palacios et al., 2021; Sadoff et al., 2021; Voysey et al., 2021), with mRNA vaccines yielding the highest efficacy in symptomatic COVID-19 prevention, i.e., 94.1 % and 95% for mRNA-1273 and BNT162b2, respectively (Baden et al., 2020; Polack et al., 2020). However, all trials have used placebo as a comparator, and there have been no head-to-head comparisons.

This study aims to demonstrate how to compare among treatment options when there is no existing direct comparison. Using the principle of network meta-analysis, the common placebo arms can facilitate indirect comparisons. Given that virtually all trials reported Kaplan-Meier (KM) curves comparing active vaccines with placebo, data from the survival curves, i.e., probability of COVID-19 infections on the y-axis and time from vaccination to infection on the x-axis, can be extracted to simulate individual participant (IP) time-to-event data (Guyot et al., 2012). Therefore, this indirect comparison was conducted to estimate the relative efficacies of the various vaccines, both individually and by class. This study is motivated by Guyot et al.’s work (Guyot et al., 2012).

## 2. Methods

Eight Phase-III randomized controlled trials (RCTs) were identified from searches of PubMed, Scopus, unpublished reports, and local FDA reports in May 2021, including CoronaVac (Sinovac), mRNA-1273 (Moderna), BNT162b2 (Pfizer BioNTech), and ChAdOx1 nCoV-19 (Astra Zeneca (AZ)/Oxford), NVX-CoV2373 (Novavax), Ad26.COV2.S (Johnson & Johnson), rAd26/rAd5 (Gamaleya), and WIV04 and HB02 (Sinopharm) (searched in May 2021). RCTs were included in this analysis if they compared time to infection and provided KM curves by intervention groups. Data including the study site, enrollment period, number of participants and events, participant characteristics, and the endpoint, were extracted.

The probability of symptomatic infection and time to infection were extracted by two independent reviewers (AT and SO) from the KM curves using WebPlotDigitizer version 4.2. In addition, the number of participants at risk, and infections at each distinct time point on the KM curve, were also extracted if available. IP data were then reconstructed from extracted data incorporating the number of participants at risk and events recorded using an algorithm reported by Wei et al (Wei & Royston, 2017). IP data for all RCTs were then combined for further analysis.

A mixed-effect accelerated failure model was applied, considering the individual study as a random effect. Survival distributions with Weibull and log-logistic models were applied where appropriate. Failure curves by vaccine type were constructed accordingly. In addition, vaccines were also grouped based on their class (i.e., inactivated virus, mRNA, and viral vector). Hazard ratios (HR) and confidence intervals (CI) were estimated to compare the risk of infection between vaccine groups. All analyses were performed using Stata version 16 (StataCrop, Texas, USA). Simulated IP data and Stata command for a mixed-effect failure model are available in Appendix 1 and 2, respectively.

## 3. Results

KM curves were available for data extraction from seven published RCTs (Al Kaabi et al., 2021; Baden et al., 2020; Logunov et al., 2021; Palacios et al., 2021; Polack et al., 2020; Sadoff et al., 2021; Voysey et al., 2021), including five (Baden et al., 2020; Logunov et al., 2021; Palacios et al., 2021; Polack et al., 2020; Sadoff et al., 2021) where the KM curves were based on an intention to treat analysis. Characteristics of the RCTs included are provided in Table 1. The seven RCTs included 201,711 participants, of which 2,265 had symptomatic COVID-19 according to the data reported in the original RCTs. The corresponding simulated data based on the KM curves included 201,711 participants, of which 2,212 had symptomatic COVID-19. The simulation was a good fit to the actual data: the discrepancies between reported and simulated events were 126 vs 120 and 252 vs 242 in vaccine and placebo groups for CoronaVac study (Palacios et al., 2021); and 79 vs 67 and 96 vs 76 in vaccine and placebo groups for rAd26/rAd5 study (Logunov et al., 2021).

**Table 1.** Trial characteristics

|  | Inactivated virus |  |  | mRNA |  | Viral vector |  |  |
| --- | --- | --- | --- | --- | --- | --- | --- | --- |
|  | CoronaVac | WIV04 | HB02 | mRNA-1273 | BNT162b2 | ChAdOx1 nCoV-19 | Ad26.COV2.S | rAd26/rAd5 |
| Study site | Brazil | UAE, Bahrain |  | US | US, Argentina, Brazil, South Africa | UK, Brazil, South Africa | US, Latin America, South Africa | Russia |
| Enrollment period | July – Dec 2020 | N/A |  | July – Oct 2020 | July – Nov 2020 | April – Nov 2020 | N/A | Sep – Nov 2020 |
| Participant characteristics |  |  |  |  |  |  |  |  |
| Age range | 18-59 | 18-60 |  | 18-65 | 16-55 | 18-55 | 18-59 | 18-60 |
| Sex, male (%) | 35.8 | 84.4 |  | 52.7 | 50.6 | 39.5 | 54.9 | 61.2 |
| Ethnicity (%) |  |  |  |  |  |  |  |  |
| White | 75.3 | N/A |  | 79.2 | 82.9 | 82.7 | 62.1 | 98.5 |
| Black | 5.2 | N/A |  | 10.2 | 9.4 | 4.1 | 17.2 | - |
| Asian | 2.5 | N/A |  | 4.6 | 4.3 | 4.4 | 3.5 | 1.4 |
| Comorbidity (%) | 55.9 | N/A |  | N/A | 21 | N/A | 40.8 | 24.8 |
| Pulmonary | N/A | N/A |  | 4.8 | N/A | 11.7 | N/A | N/A |
| Cardiac | 12.6 | N/A |  | 4.9 | N/A | 10.7 | N/A | N/A |
| Obesity | 22.5 | N/A |  | 6.7 | N/A | N/A | N/A | N/A |
| Endpoint | Symptomatic COVID-19 after the first vaccination (ITT) | Symptomatic COVID-19 after 2 weeks of the second vaccination (PP) |  | Symptomatic COVID-19 after randomization (ITT) | Symptomatic COVID-19 after the first vaccination (ITT) | Symptomatic COVID-19 after 2 weeks of the second vaccination (PP) | Moderate to severe COVID-19 with onset at least 1 day after vaccination (ITT) | Symptomatic COVID-19 after the first vaccination (ITT) |
| Participants: Actual/simulated N |  |  |  |  |  |  |  |  |
| Vaccine | 6195/6195 | 12743/12743 | 12726/12726 | 14550/14550 | 21669/21669 | 8597/8597 | 19744/19744 | 16427/16427 |
| Placebo | 6201/6201 | 12737/12737 |  | 14598/14598 | 21686/21686 | 8581/8581 | 19822/19822 | 5435/5435 |
| Events: Actual/simulated N |  |  |  |  |  |  |  |  |
| Vaccine | 126/120 | 26/25 | 21/21 | 19/18 | 50/50 | 84/83 | 193/192 | 79/67 |
| Placebo | 252/242 | 95/94 |  | 269/269 | 275/275 | 248/249 | 432/431 | 96/76 |

N/A not available, PP per protocol, ITT intention to treat

A mixed-effect accelerated failure model with log-logistic distribution was used to estimate the probability of infection. All vaccines had greater efficacy compared to placebo, but the highest probability of infection was for CoronaVac, followed by Ad26.COV2.S, ChAdOx1 nCoV-19, rAd26/rAd5, WIV04, HB02, BNT162b2, and mRNA-1273 (see Figure 1). The HRs (95% CI) were 0.07 (0.04, 0.11), 0.14 (0.10, 0.19), 0.21 (0.13, 0.33), 0.25 (0.16, 0.39), 0.29 (0.21, 0.41), 0.33 (0.25, 0.42), 0.44 (0.37, 0.52), and 0.50 (0.40, 0.62) for mRNA-1273, BNT162b2, HB02, WIV04, rAd26/rAd5, ChAdOx1 nCoV-19, Ad26.COV2.S, and CoronaVac versus placebo, respectively, see Table 2. Comparing vaccines, CoronaVac, Ad26.COV2.S, ChAdOx1 nCoV-19, rAd26/rAd5, WIV04, HB02, and BNT162b2 showed 7.61 (4.50, 12.87), 6.77 (4.08, 11.24), 5.01 (2.93, 8.57), 4.50 (2.52, 8.01), 3.90 (2.04, 7.45), 3.18 (1.62, 6.21), and 2.15 (1.22, 3.78) times significantly higher risk of infection than mRNA-1273. CoronaVac, Ad26.COV2.S, ChAdOx1 nCoV-19, rAd26/rAd5, WIV04, and HB02 showed 3.54 (2.44, 5.14), 3.15 (2.23, 4.45), 2.33 (1.58, 3.44), 2.09 (1.34, 3.26), 1.81 (1.07, 3.09), and 1.48 (0.85, 2.58) times higher risk of infection than BNT162b2.

**Figure 1.**
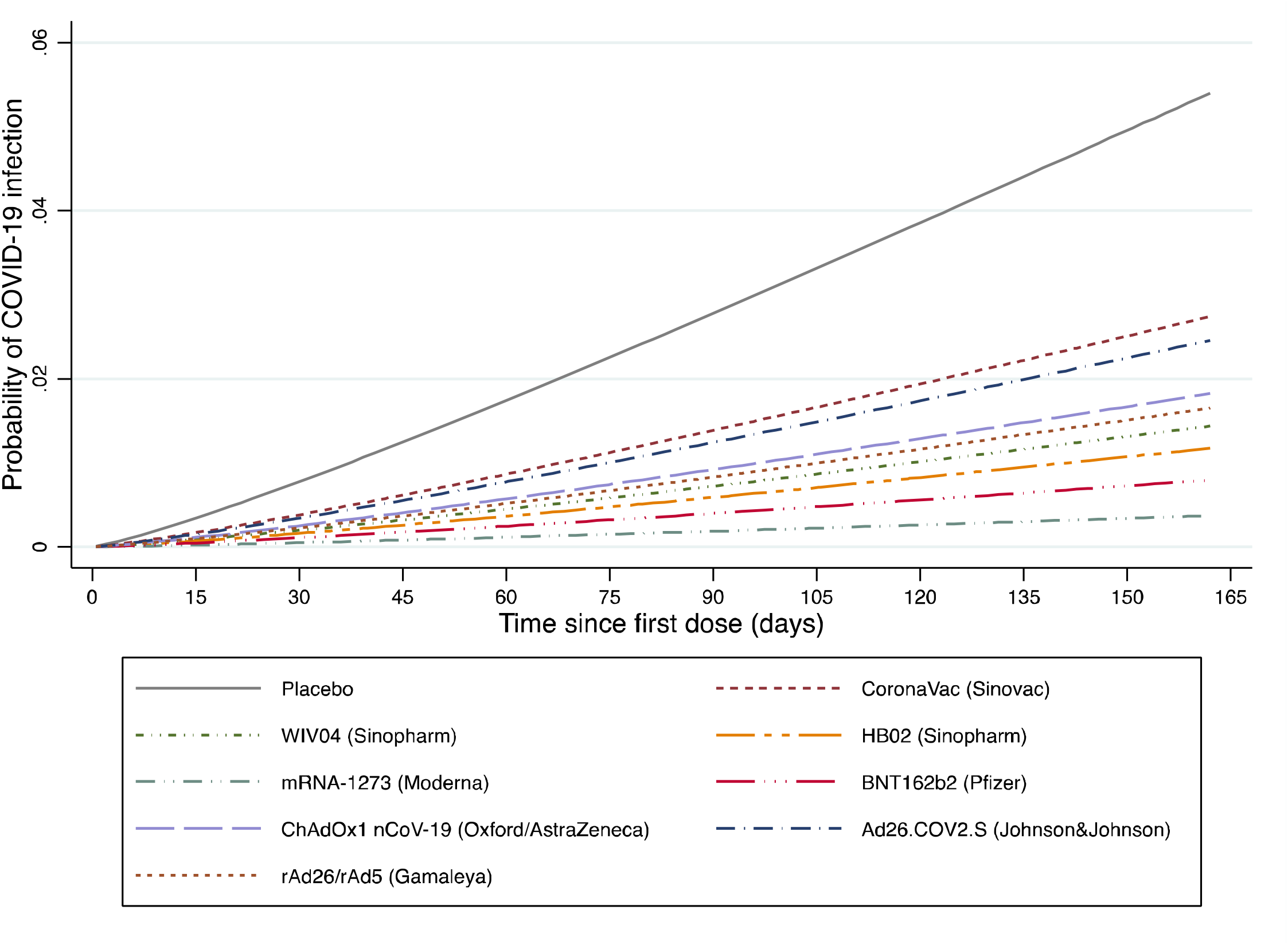
Cumulative incidence of Covid-19 infection curves by individual vaccine

**Table 2.**
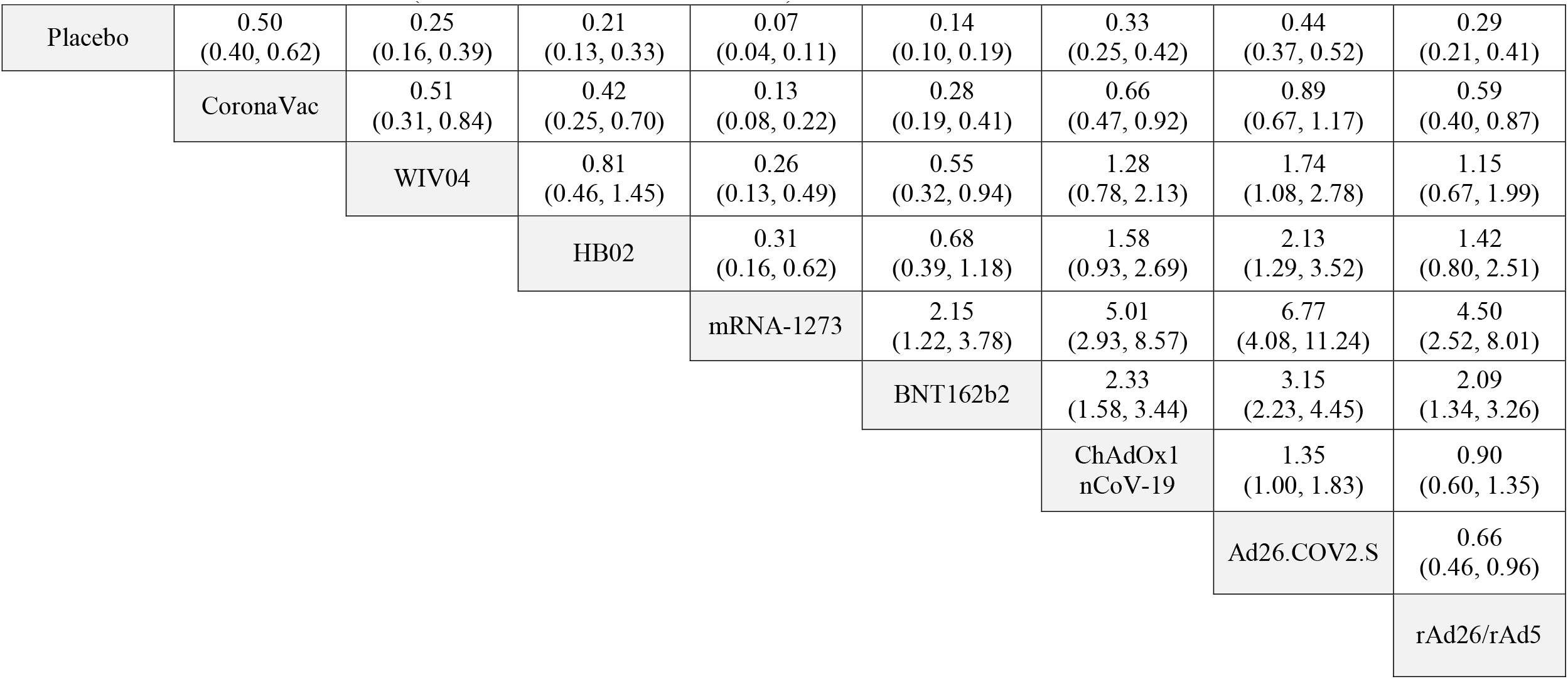
Hazard ratio estimates (and 95% confidence intervals) of relative COVID-19 vaccine effects.

When considered as classes, the mRNA vaccines yielded a significantly lower risk of infection, followed by the viral vector and inactivated viral vaccines (see Figure 2) with HRs of 0.11 (0.08, 0.14), 0.38 (0.33, 0.43), and 0.39 (0.33, 0.47), respectively, see Table 3. In addition, mRNA and viral vector vaccines showed a lower risk of infection compared with the inactivated viral vaccine but the later was not significant, with HRs (95% CI) of 0.27 (0.20, 0.37) and 0.97 (0.77, 1.22), respectively.

**Table 3.**
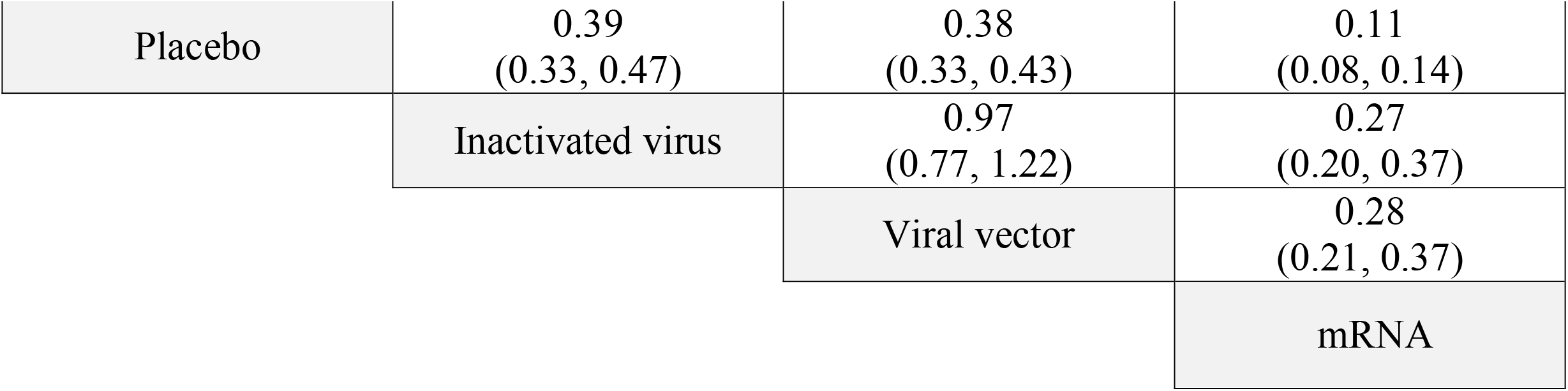
Hazard ratio estimates (and 95% confidence intervals) of relative COVID-19 vaccine effects by class.

|  |  |  |  |
| --- | --- | --- | --- |
| Placebo | 0.39<br>(0.33, 0.47) | 0.38<br>(0.33, 0.43) | 0.11<br>(0.08, 0.14) |
|  | Inactivated virus | 0.97<br>(0.77, 1.22) | 0.27<br>(0.20, 0.37) |
|  |  | Viral vector | 0.28<br>(0.21, 0.37) |
|  |  |  | mRNA |

**Figure 2.**
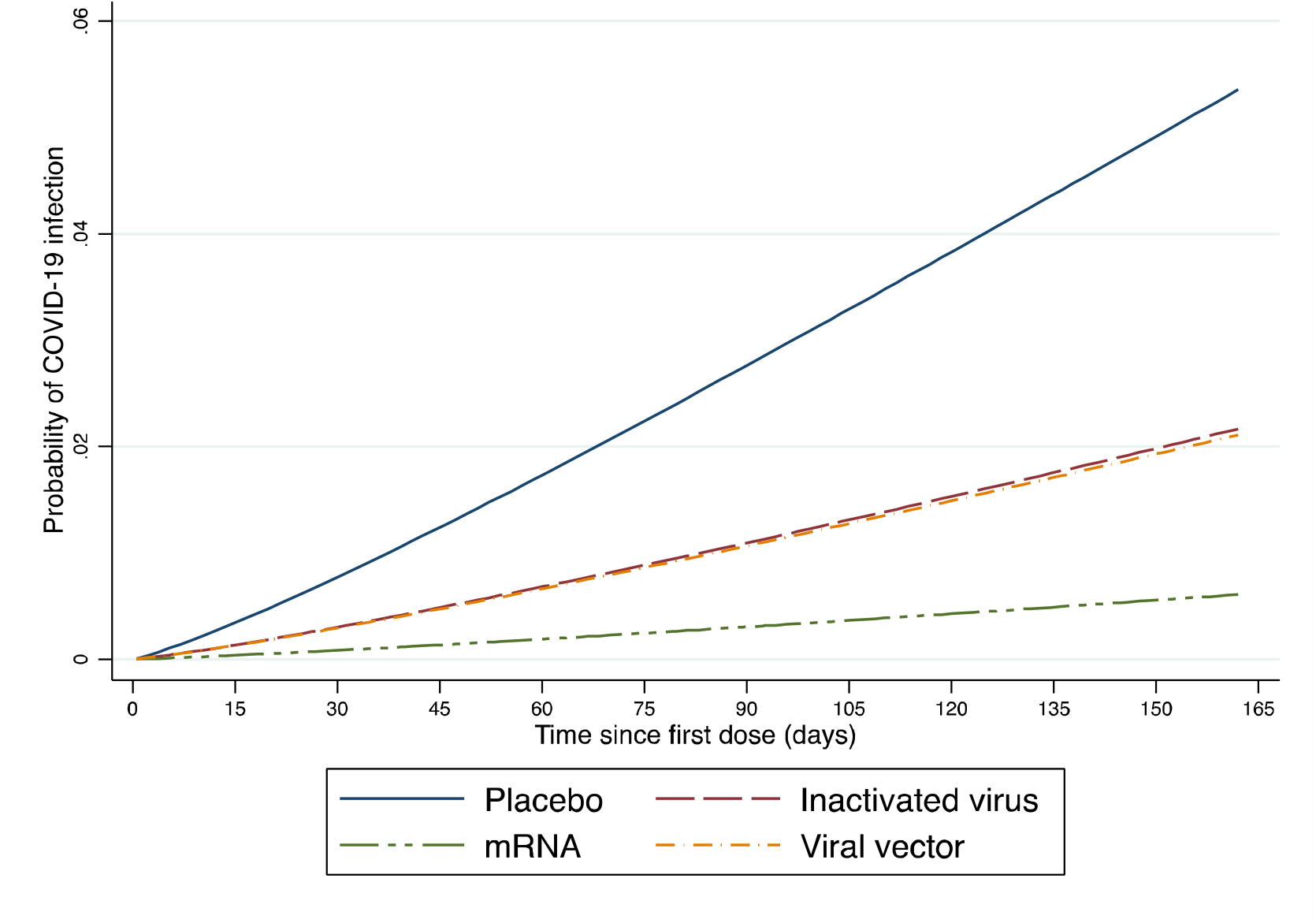
Cumulative incidence of Covid-19 infection curves by vaccine class

## Discussion and Conclusion

Data from KM curves were extracted from seven RCTs that assessed the efficacy of various COVID-19 vaccines compared to placebo, and IP time to event data were simulated. Merging IP data from each trial allowed further analysis using a mixed-effect model. The mixed-effect model indicated that all vaccines had significant protective effects compared to no vaccination. All vaccines, with the exception of Ad26.COV2.S, also had significantly better efficacy than CoronaVac, with mRNA-1273 vaccine showing the highest efficacy. As a class, mRNA vaccines (mRNA-1273 and BNT162b2) were more efficacious than viral vector and inactivated virus vaccines in preventing symptomatic COVID-19.

At the beginning of pandemic, data were limited, whereas comparison among the vaccines’ efficacy was needed. Actually, no RCT has directly compared vaccine efficacy until present. This study aims to demonstrate how to address this problem which could guide national vaccine policy. However, there are some limitations. First, infection rates were indirectly compared between vaccines using placebo as a common comparator. The RCTs included were conducted in different settings where baseline risks of infection varied across studies, ranging from 0.7% to 3.9%. Second, trials included in this study have different endpoint (see Table 1) and results might be biased from this fact. Finally, simulating IP data from KM curves resulted in some variation in the number of events between original reports and the simulated data, although this discrepancy was small except for two RCTs (Logunov et al., 2021; Palacios et al., 2021).

In conclusion, combining IP data simulation from KM curves and mixed-effect accelerated failure modeling allows us to compare COVID-19 vaccine’s efficacy when there is no head-to-head comparison. This evidence indicates that although all vaccines showed strong efficacy in reducing COVID-19 infection compared to no vaccination, there are small but significant differences between them, with mRNA vaccines showing the highest efficacy, followed by viral vector and inactivated viral vaccines.

## Data Availability

All data produced in the present study are available upon reasonable request to the authors

## Author contribution

Amarit Tansawet, Songporn Oranratnachai, and Grace Filbertine contributed to the search of original articles, appraised evidence, and extracted data. Statistical analysis was performed by Amarit Tansawet, Songporn Oranratnachai, Pawin Numthavaj, and Ammarin Thakkinstian. Manuscript was drafted by Amarit Tansawet, and revised by Sasisopin Kiertiburanakul, Gareth J McKay, John Attia, and Ammarin Thakkinstian. All submitted materials have been approved by all authors.

## Conflict of interest

All authors declare no competing interest.

### IRB approval

Not applicable

### Funding source

This research did not receive any specific grant from funding agencies in the public, commercial, or not-for-profit sectors.

### Patient and Public involvement

None

### Dissemination declaration

Dissemination to participants is not applicable.

### Data availability statement

Simulated data are available in Appendix 1.

## Appendix

Appendix 1. Simulated individual participant data. xlsm

Appendix 2. Stata commands for a mixed-effect failure model. docx

## References

Al Kaabi, N., Zhang, Y., Xia, S., Yang, Y., Al Qahtani, M. M., Abdulrazzaq, N., … Yang, X. (2021). Effect of 2 Inactivated SARS-CoV-2 Vaccines on Symptomatic COVID-19 Infection in Adults: A Randomized Clinical Trial. JAMA, 326(1), 35–45. doi:10.1001/jama.2021.8565

Baden, L. R., El Sahly, H. M., Essink, B., Kotloff, K., Frey, S., Novak, R., … Zaks, T. (2020). Efficacy and Safety of the mRNA-1273 SARS-CoV-2 Vaccine. New England Journal of Medicine, 384(5), 403–416. doi:10.1056/NEJMoa2035389

Guyot, P., Ades, A. E., Ouwens, M. J., & Welton, N. J. (2012). Enhanced secondary analysis of survival data: reconstructing the data from published Kaplan-Meier survival curves. BMC Medical Research Methodology, 12, 9. doi:10.1186/1471-2288-12-9

Logunov, D. Y., Dolzhikova, I. V., Shcheblyakov, D. V., Tukhvatulin, A. I., Zubkova, O. V., Dzharullaeva, A. S., … Gintsburg, A. L. (2021). Safety and efficacy of an rAd26 and rAd5 vector-based heterologous prime-boost COVID-19 vaccine: an interim analysis of a randomised controlled phase 3 trial in Russia. The Lancet, 397(10275), 671–681. doi:10.1016/S0140-6736(21)00234-8

World Health Organization. (2021). WHO Coronavirus (COVID-19) Dashboard. Retrieved from https://covid19.who.int.

Palacios, R., Batista, A. P., Albuquerque, C. S. N., Patiño, E. G., Santos, J. d. P., Conde, M. T. R. P., … Kallas, E. G. (2021). Efficacy and safety of a COVID-19 Inactivated Vaccine in Healthcare Professionals in Brazil: The PROFISCOV Study.. doi:https://dx.doi.org/10.2139/ssrn.3822780

Polack, F. P., Thomas, S. J., Kitchin, N., Absalon, J., Gurtman, A., Lockhart, S., … Gruber, W. C. (2020). Safety and Efficacy of the BNT162b2 mRNA Covid-19 Vaccine. New England Journal of Medicine, 383(27), 2603–2615. doi:10.1056/NEJMoa2034577

Sadoff, J., Gray, G., Vandebosch, A., Cárdenas, V., Shukarev, G., Grinsztejn, B., … Douoguih, M. (2021). Safety and Efficacy of Single-Dose Ad26.COV2.S Vaccine against Covid-19. New England Journal of Medicine. doi:10.1056/NEJMoa2101544

Voysey, M., Costa Clemens, S. A., Madhi, S. A., Weckx, L. Y., Folegatti, P. M., Aley, P. K., … Zuidewind, P. (2021). Single-dose administration and the influence of the timing of the booster dose on immunogenicity and efficacy of ChAdOx1 nCoV-19 (AZD1222) vaccine: a pooled analysis of four randomised trials. The Lancet, 397(10277), 881–891. doi:10.1016/S0140-6736(21)00432-3

Wei, Y., & Royston, P. (2017). Reconstructing time-to-event data from published Kaplan-Meier curves. Stata Journal, 17(4), 786–802.

